# Epileptic encephalopathy linked to a DALRD3 missense variant impaired in tRNA modification function

**DOI:** 10.1101/2024.05.21.24307194

**Authors:** Kejia Zhang, Katharina Löhner, Henny H. Lemmink, Maartje Boon, Jenna M. Lentini, Naduni de Silva, Dragony Fu

## Abstract

Epileptic encephalopathies are severe epilepsy syndromes characterized by early onset and progressive cerebral dysfunction. A nonsense variant in the *DALR Anticodon Binding Domain Containing 3* (*DALRD3*) gene has been implicated in epileptic encephalopathy but no other disease-associated variants in *DALRD3* have been described. In human cells, the DALRD3 protein forms a complex with the METTL2 methyltransferase to generate the 3-methylcytosine (m3C) modification in specific arginine tRNAs. Here, we identify a patient with a homozygous missense variant in DALRD3 who displays developmental delay, cognitive deficiencies, and multifocal epilepsy. The missense variant substitutes an arginine residue to cysteine (R517C) within the DALR domain of DALRD3 that is required for binding tRNAs. Patient cells homozygous for the DALRD3-R517C variant exhibit reduced levels of m3C modification in arginine tRNAs, indicating that the R517C variant impairs DALRD3 function. Notably, the DALRD3-R517C variant displays reduced association with METTL2 and loss of interaction with substrate tRNAs. Our results uncover a novel pathogenic variant in DALRD3 that links DALRD3 loss-of-function to epileptic encephalopathy disorders. Importantly, these findings underscore DALRD3-dependent tRNA modification as a key contributor to proper brain development and function.

## Main Text

The post-transcriptional modification of transfer RNA (tRNA) has emerged as a critical modulator of biological processes ranging from gene expression to development^1,2^. In particular, tRNA modifications play critical roles in tRNA structure, stability, and function in protein synthesis^3-5^. Defects in tRNA modification have emerged as the cause for many types of human diseases, highlighting the critical role of tRNA modification in human health and physiology^6-8^. Notably, the brain appears to be sensitive to perturbations in tRNA modifications, as evidenced by the numerous neurological and neurodevelopmental disorders linked to deficiencies in tRNA modification^9,10^.

In mammalian cells, a subset of arginine tRNAs contain the 3-methylcytosine (m3C) modification at position 32 of the anticodon loop^11,12^. The m3C modification is hypothesized to stabilize the folding of the anticodon loop and has been shown to influence mitochondrial tRNA structure in human cells^13,14^. Thus, the m3C modification could affect the function of tRNAs in translation and protein expression (reviewed in ^15^). In human cells, the METTL2 methyltransferase forms a complex with the DALR Anticodon Binding Domain Containing 3 (DALRD3) protein to methylate tRNA-Arg-UCU and tRNA-Arg-CCU^16^. The DALRD3 protein recognizes specific arginine-tRNA isoacceptors to target them for methylation by the METTL2 methyltransferase. Human cells deficient in DALRD3 exhibit nearly complete loss of m3C in tRNA-Arg-UCU and Arg-CCU, demonstrating that DALRD3 plays a key role in m3C formation in specific arginine tRNAs^16^.

We have previously identified two sibling patients with a homozygous nonsense variant in exon 9 of the DALRD3 gene (rs1163930676, NM_001276405.1:c.1251 C > A, p.(Tyr417*))^16^. The C to A transversion introduces a premature stop codon in the DALRD3 mRNA and results in the loss of DALRD3 protein expression. The two patients were born from consanguineous parents who are heterozygous for the nonsense variant. The parents are healthy and do not exhibit any detectable pathologies. In contrast, the homozygous siblings display a collection of clinical symptoms classified as severe developmental and epileptic encephalopathy disorder^17^. Patient cells from the affected siblings exhibit a substantial reduction in m3C modification in tRNA-Arg-CCU and tRNA-Arg-UCU. These findings suggest a crucial biological role for DALRD3-dependent tRNA modification in development and nervous system function. However, the DALRD3 nonsense allele represents the only case of a DARLD3 variant that has been implicated in a neurodevelopmental disorder.

Here, we describe a human patient exhibiting severe developmental delay and multi-focal epilepsy associated with a novel variant in *DALRD3* (Table 1, patient 1). The patient was born at term via Cesarian section. Seizures started after a febrile episode followed by regression of psychomotor skills. The patient was referred to a pediatric neurologist for analysis because of progression of his symptoms. At neurological examination the patient displayed a dropped head and reduced facial expressivity (Figure 1A). In addition, he had progressive immobility, limited speech consisting of one-word expressions, increased muscle tone in his arms, subtle myoclonus and ataxia.

**Table 1.** Clinical phenotype of patients with homozygous variants in DALRD3.

| **Patient** | 1 (this study) | 2 (Lentini et al. 2020) | 3 (Lentini et al. 2020) |
| --- | --- | --- | --- |
| **Genotype** | NM_001009996.2: c.1549C>T, homozygous | NM_001276405.1:c.1251C>A: | NM_001276405.1:c.1251C>A: |
| **Protein** | p.(Arg517Cys) | p.(Tyr417*) | p.(Tyr417*) |
| **Perinatal history** | Uncomplicated pregnancy. Born term by caesarean section due to previous caesarean section. | Normal spontaneous vaginal delivery with history of placental insufficiency and oligohydramnios | Full term product of caesarean section due to breech presentation and oligohydramnios in addition to placental insufficiency |
| **Weight at birth (kg)** | Unknown | 2.25 (-2.2 SD) | 2.5 (4<sup>th</sup> centile) |
| **Developmental delay** | Severe | Severe | Severe |
| **Motor** | Progressively immobile, can walk some steps unaided | Immobile | Immobile |
| **Seizures** | Seizures started at childhood after febrile episode. Thereafter regression of psychomotor skills. Focal clonic seizures with impaired awareness to bilateral seizures, often in clusters up to 10, during the night /early morning. As teenager, interictal subtle myoclonus of the hands and fingers is seen. | Seizures started at infancy in the form of myoclonic jerks which remains frequent and poorly controlled by antiepileptic medications | Epilepsy ensued at infancy, initially as brief episodes of flexion tonic spasm of head followed by myoclonic seizures. Unlike the sibling, the epilepsy of patient 2 is reasonably controlled by antiepileptic medications. |
| **EEG** | Intermittent focal epileptic phenomena in temporal regions, slow background activity without normal differentiation, high amplitude slow waves over frontotemporal regions. | Independent multifocal epileptic discharges predominantly over the anterior head region bilaterally as well as over the right temporal and right parietal regions | Markedly high voltage and slow background for age along with slow generalized polyspike and wave activity |
| **Tone** | Axial hypotonic, extremities slightly hypertonic. Initially higher, later low tendon reflexes with extensor plantar responses. Some ataxia of gait and hands. | Axial and peripheral hypotonia with dystonic like movement and generalized muscle wasting | Central and peripheral hypotonia with dystonic like movements and generalized muscle wasting |
| **Microcephaly** | No | No | Yes |
| **Brain finding** | Enlargement of central and peripheral CSF spaces. Normal aspect of white matter, basal ganglia, thalamus, and brainstem. Arteriovenous malformation in right cerebellar hemisphere, otherwise normal cerebellum. Thickened skull by broad diploic space. | Mild diffuse brain parenchymal volume loss with diffuse paucity of the myelin within the brain parenchyma. | Normal topographical and morphological appearance of the infratentorial and supratentorial structures |
| **Audiology assessment** | N/A | Moderate to severe conductive hearing loss in the left ear and mild conductive hearing loss in right ear | N/A |

**Figure 1.**
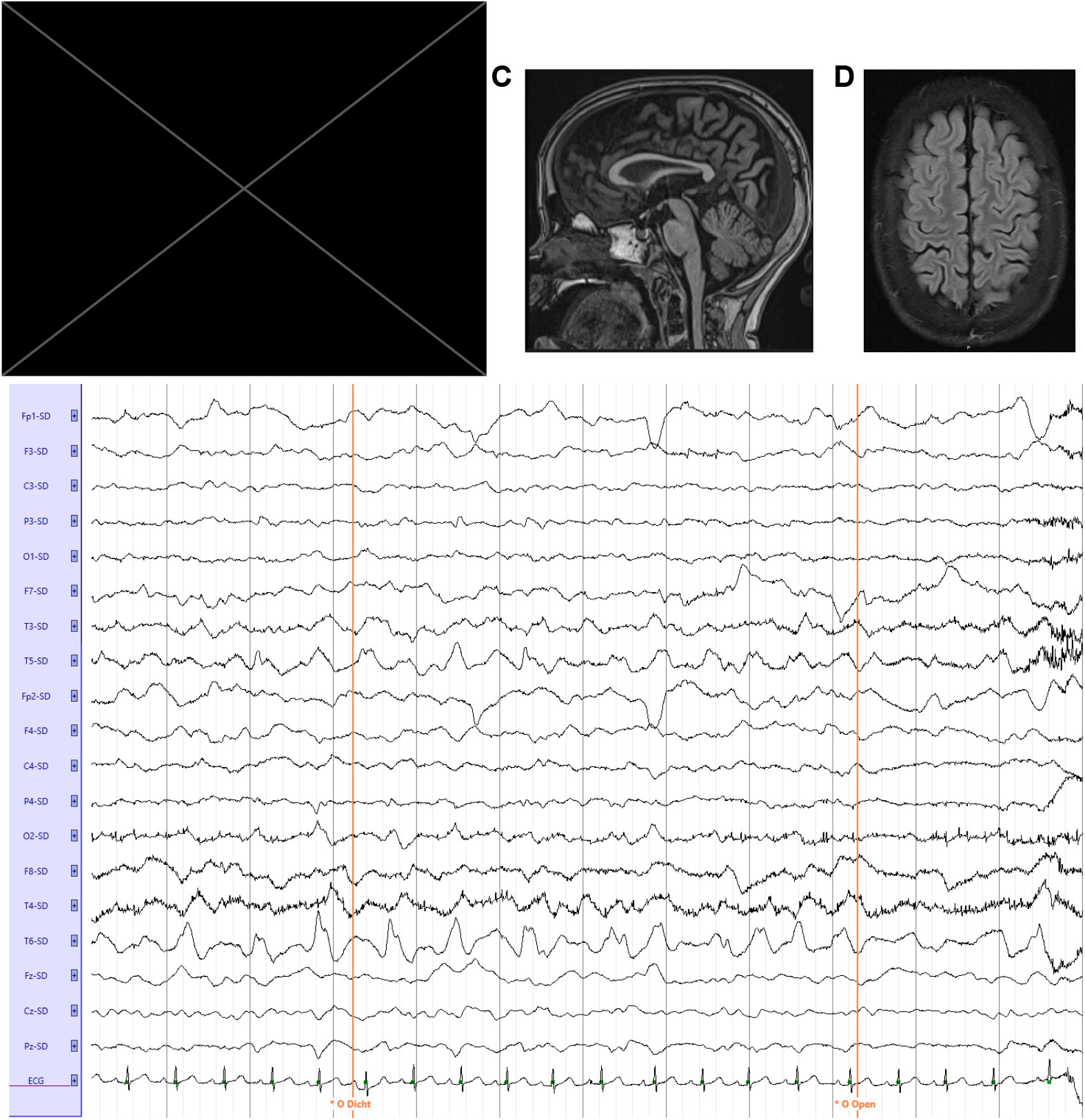
Identification of an individual exhibiting epileptic encephalopathy and brain pathologies. (A) Patient containing a homozygous c.1549C>T (NM_001009996.3) variant in *DALRD3*. (B) EEG trace, Laplacian montage with eyes closed and opened. (C) MRI of the patient, Sagittal T1 MPRAGE. (D) MRI of the patient, Transversal T2 TSE.

EEG testing revealed slow background activity without normal differentiation, intermittent focal epileptic phenomena in temporal regions, and high amplitude slow waves over frontotemporal regions (Figure 1B, Supplemental Figure 1). No microcephaly was detected, but there was relatively thin corpus callosum, enlargement of central and peripheral CSF spaces, along with an arteriovenous malformation in the right cerebellar hemisphere (Figure 1C and 1D, Supplemental Figure 2). Based upon the pattern and onset of symptoms, the patient matches clinical conditions classified as epileptic encephalopathy^17^.

Exome sequencing in the affected patient revealed the presence of a homozygous nucleotide substitution resulting in a C to T transition in the last exon of the *DALRD3* gene (Figure 2A, Supplemental Figure 3). Sanger sequencing in a healthy, control individual and the patient confirmed the homozygous nucleotide substitution in the affected patient (Figure 2B). The substitution causes a missense variant in the encoded DALRD3 protein of the canonical transcript (NM_001009996.3: c.1549C>T, p(Arg517Cys)). Translation of this transcript is expected to produce a protein containing an arginine to cysteine substitution at position 517 (R517C) in the DALR tRNA anticodon-binding domain (Figure 2A, protein). The parents of the patient are consanguineous and heterozygous carriers of the DALRD3 variant (Figure 2C, Supplemental Figure 4). Both parents are healthy with normal development and no detectable epilepsy. The DALRD3 variant was not detected in the patient’s older sibling, who is healthy, developed normally, and did not exhibit epilepsy. The genotype of the other sibling is unknown, but she is healthy and exhibits normal development. These studies suggest that the R517C missense variant segregates with the disease in the family in an autosomal-recessive manner, being heterozygous in the parents, absent in the healthy sibling, and homozygous in the affected patient.

**Figure 2.**
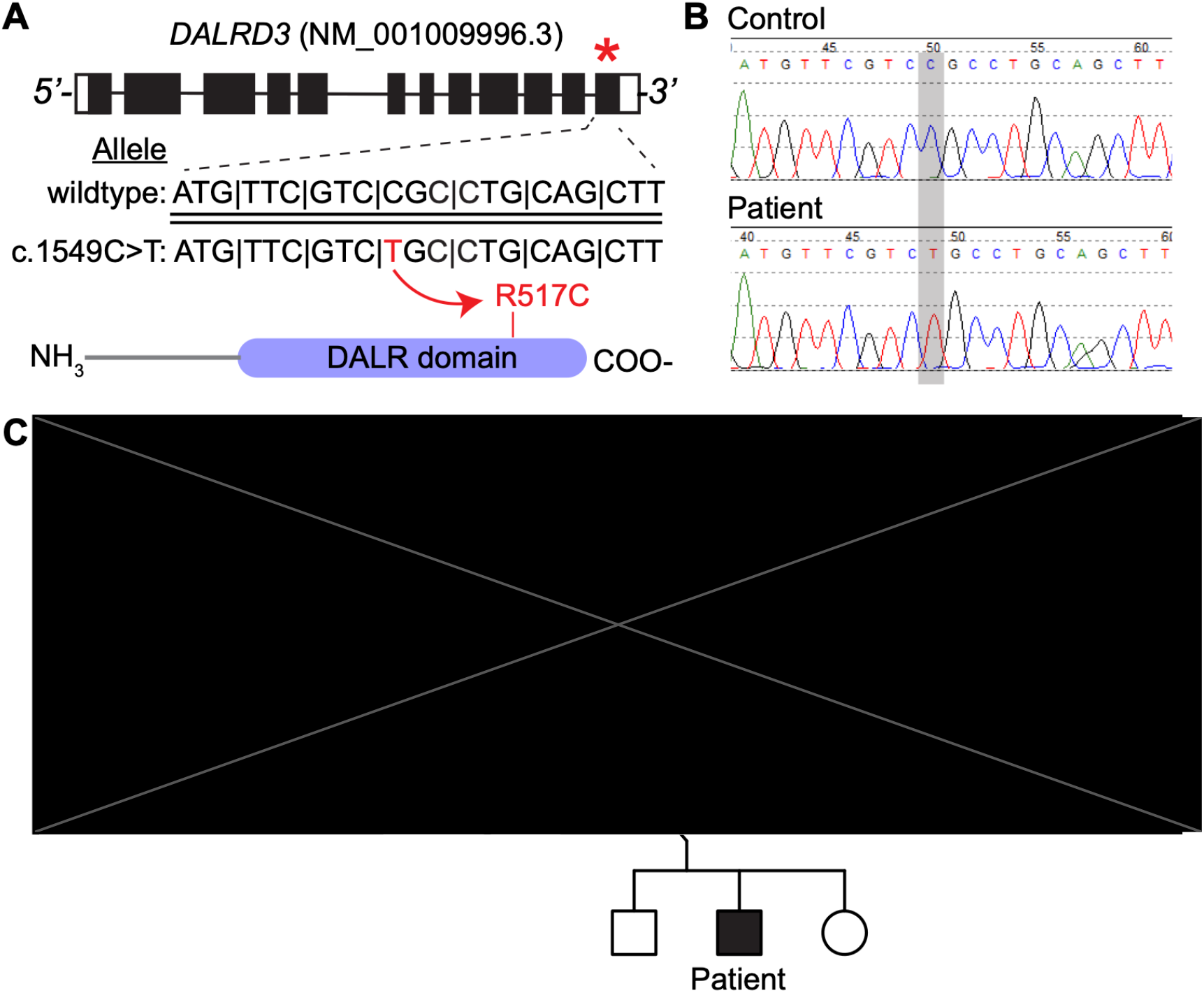
Identification of a novel missense variant in the DALRD3 gene associated with epileptic encephalopathy. (A) *DALRD3* exon and intron structure with encoded protein shown below. Asterisk represents location of the variant in the mRNA (NM_001009996.3). Wildtype and c.1549C>T alleles are shown with the variant shown in red. (B) Sanger sequencing chromatograms of a healthy, control individual and the patient characterized in this study. (C) Pedigree of the family harboring the missense variant in the *DALRD3* gene.

Sequence alignment reveals that the R517 residue of human DALRD3 is conserved in all known vertebrate DALRD3 homologs from mammals to fish (Figure 3A). Based upon the high conservation in amino acid identity at this position, the nonsynonymous substitution caused by the R517C variant could disrupt the folding and function of the DARLD3 protein. Consistent with this hypothesis, the R517 variant is expected to be deleterious by multiple pathogenicity prediction algorithms (Figure 3B).

**Figure 3.**
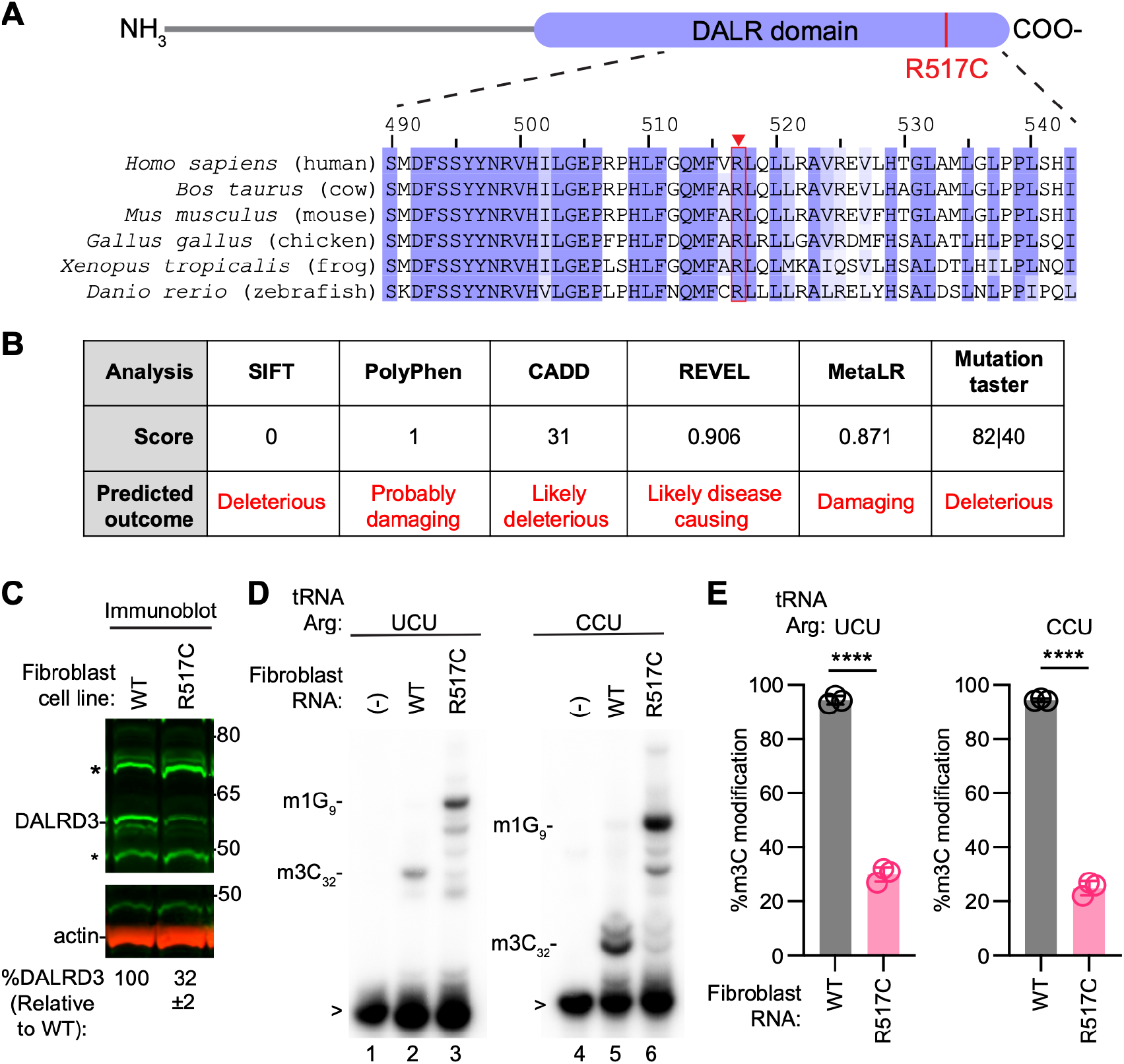
Patient cells homozygous for the R517C variant exhibit decreased levels of DALRD3 protein and a reduction in m3C modification in arginine tRNAs. (A) Schematic of the DALRD3 protein and alignment of the region encompassing the R517C variant. (B) Scores and predicted outcomes for the R517C variant based upon the indicated pathogenicity prediction algorithms. (C) Immunoblot of lysates prepared from wildtype or R517 fibroblast cell lines. %DALRD3 represents the amount of DALRD3 protein relative to the WT fibroblast cell line and was quantified from three independent samples. ±, standard deviation from the mean. (D) Primer extension analysis of tRNA-Arg-UCU and Arg-CCU extracted from wildtype or R517 fibroblast cell lines. (-) represents no RNA was added to the RT reaction. m3C_32_-3-Methylcytosine. m1G_9_- 1-methylguanine; > labelled probe. (E) Quantification of m3C formation in tRNA-Arg-UCU or CCU by primer extension. n = 3. Error bars represent standard deviation from the mean. Significance was determined using an unpaired t-test with two-tailed P value. ****P ≤ 0.0001.

To characterize the biological effects of the DALRD3-R517C variant, we isolated fibroblast cells from the affected patient via skin biopsy (referred to as R517C fibroblast cells). We compared protein levels in the R517C fibroblast cells to fibroblast cells isolated from a healthy, age-matched control individual expressing wildtype DALRD3. Based upon immunoblotting, the levels of DALRD3 protein in the R517C patient fibroblast cells were greatly reduced compared to fibroblast cells from the healthy, wildtype individual (Figure 3C). These results suggest that the R517C variant alters the structure of DALRD3 leading to reduced cellular stability and increased degradation.

We next tested the impact of the DALRD3-R517 variant on m3C modification in arginine tRNAs of patient fibroblast cells. We monitored m3C modification using a primer extension assay in which the presence of m3C leads to a reverse transcriptase (RT) block at position 32 of arginine tRNAs while the lack of m3C allows for read-through and generation of an extended product to the next RT block. Using this assay, we find that the m3C modification is greatly reduced in R517C fibroblast cells compared to control cells from a healthy individual (Figure 3D, quantified in 3E). These results demonstrate that R517C patient fibroblasts exhibit a deficiency in the m3C modification in specific tRNA-Arg isoacceptors. The deficit in m3C modification in the arginine tRNAs of patient LCLs provides evidence that the DALRD3-R517C variant causes partial loss-of-function.

The above results indicate that the DALRD3-R517C variant impacts m3C modification in human cells. To elucidate the molecular defects associated with the DALRD3-R517C variant, we investigated the interaction between DALRD3 and METTL2. We co-transfected 293T human embryonic cells with a plasmid expressing Strep-tagged METTL2 along with empty vector or plasmids expressing a FLAG-tagged version of wildtype (WT) or the R517C DALRD3 variant (Figure 4A). Expression of METTL2, DALRD3-WT or the DALRD3-R517C variant was confirmed by immunoblotting for the Strep tag or FLAG tag (Figure 4B, Input, lanes 1 through 3). We note that the DALRD3-R517C variant accumulated to lower levels than the wildtype DALRD3 protein in multiple independent transfections. The reduced levels of DALRD3-R517C protein compared to WT-DALRD3 in this system is consistent with the decreased amount of DALRD3 protein in homozygous patient cells with the R517C variant.

**Figure 4.**
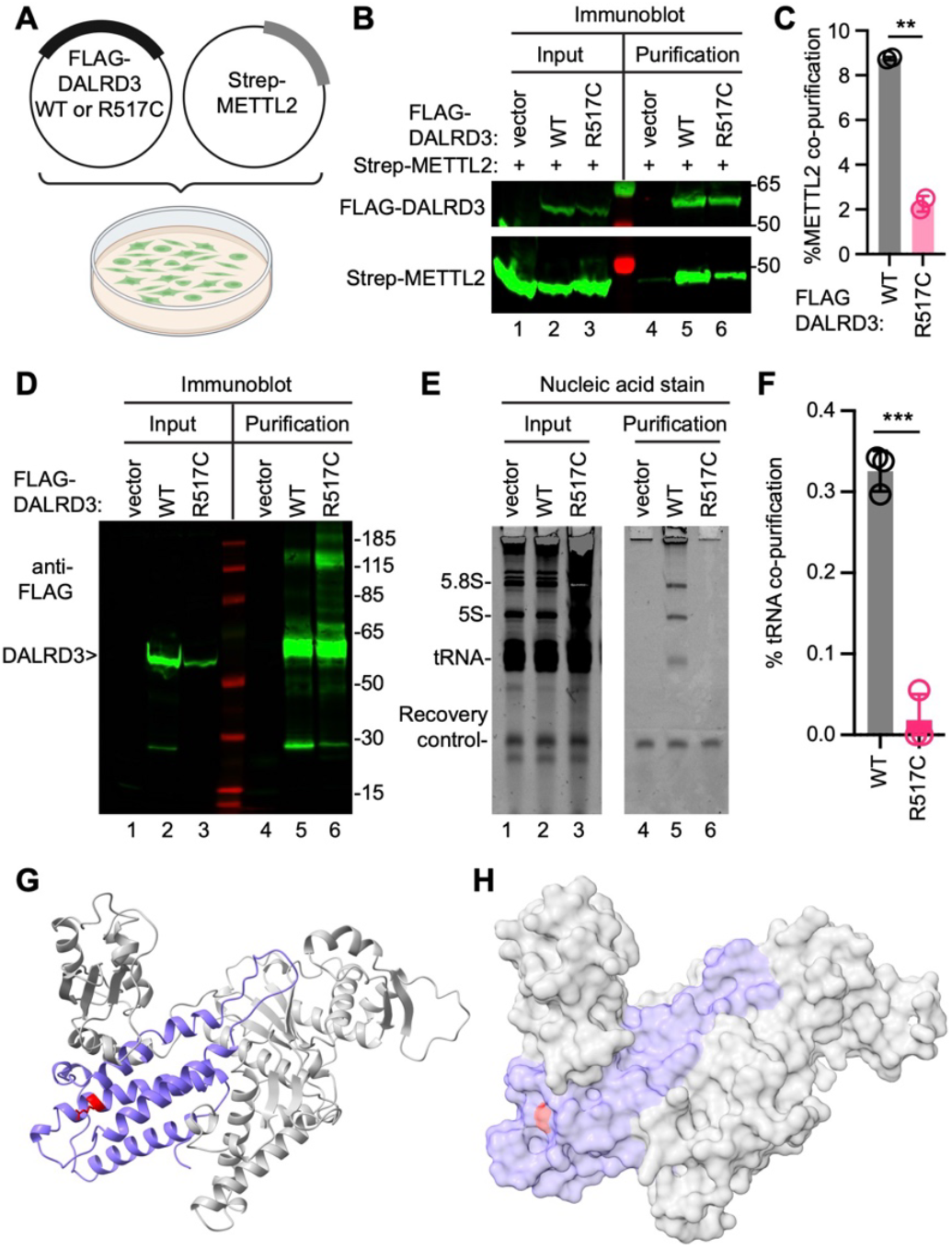
The DALRD3-R517C variant exhibits reduced co-precipitation with METTL2 and RNAs. (A) Co-transfection setup with plasmids encoding FLAG-DALRD3 and Strep-METTL2. (B) Immunoblot of input lysates and anti-FLAG purifications from 293T cells transfected with the indicated plasmids. Immunoblot was probed with anti-FLAG and anti-Strep antibodies. Input represents 4% of the total sample used for purification. Purification represents 50% of the purified sample. (C) Quantification of METTL2 copurifying with DALRD3-WT or R517C variant. The percentage of co-purifying METTL2 represents the amount of METTL2 in the purified DALRD3 sample that was recovered from the total input. (D) Immunoblot of FLAG-DALRD3 purified from 293T cells. Immunoblot was probed with anti-FLAG antibodies. Input represents 4% of the total sample used for purification. Purification represents 20% of the total purified sample. (E) Nucleic acid stain of RNAs extracted from the indicated input or purified samples after denaturing PAGE. The migration pattern of 5.8S rRNA (∼150 nt), 5S rRNA (∼120 nt) and tRNAs (∼70–80 nt) are denoted. Input represents 2% of total extracts used for purification. (F) Quantification of tRNA copurifying with DALRD3-WT or R517C variant. The percentage tRNA co-purification represents the amount of tRNA in the purified DALRD3 sample that was recovered from the total input. For (C) and (F), error bars represent standard deviation from the mean; significance was determined using an unpaired t-test with two-tailed P value. **P = 0.0015 for (C). ***P = 0.0002 for (F). (G) AlphaFold model of human DALRD3. The DALR anticodon binding domain is represented in violet and the R517 residue noted in red. (H) Surface plot of the predicted human DALRD3 structure with the R517 residue denoted in red.

The FLAG-DALRD3 fusion proteins were then purified on anti-FLAG antibody resin and recovery of FLAG-tagged DALRD3 was confirmed (Figure 4B, Purification, FLAG-DALRD3, lanes 5 and 6). We detected an enrichment of METTL2 that copurified with DALRD3 above the background binding in the control purification (Figure 4B, Purification, Strep-METTL2, compare lane 4 to lane 5). The amount of copurifying METTL2 was decreased by ∼4-fold with the DALRD3-R517C variant compared to DALRD3-WT (Figure 4B, Strep-METTL2, compare lanes 5 and 6, quantified in Figure 4C). This result suggests that the DALRD3-R517C variant exhibits less stable interaction with METTL2 compared to wildtype DALRD3.

Based upon this finding, we next tested the interaction of DALRD3 with tRNAs. Like above, we transfected 293T human embryonic cells with empty vector or plasmids expressing FLAG-DALRD3-WT or the DALRD3-R517C variant. Expression and purification of DALRD3-WT or the DALRD3-R517C variant was confirmed by immunoblotting for the FLAG tag (Figure 4D). We examined the RNA species that co-purified with DALRD3 by denaturing PAGE followed by nucleic acid staining. A recovery control RNA was included during RNA extraction to normalize for differences in recovery efficiency. While purification from vector-transfected cell lysates exhibited only background binding to tRNAs, the purified DALRD3-WT sample contained several co-purifying RNA species that correspond in size to tRNAs along with 5S and 5.8S rRNA (Figure 4E, compare lanes 4 to 5). This pattern of copurifying RNAs is consistent with our previous observation that DALRD3 exhibits a stable interaction with rRNA and tRNAs that are targets for m3C modification^16^. In contrast to DALRD3-WT, the amount of copurifying tRNAs with the DALRD3-R517C variant was reduced to nearly background levels (Figure 4E, quantified in 4F). Overall, these results reveal that the R517C missense variant perturbs binding of DALRD3 to METTL2 and tRNAs.

To gain insight into how the R517C variant could affect interaction with METTL2 and tRNA substrates, we mapped the R517 residue onto a predicted DALRD3 structure generated through AlphaFold^18,19^. DALRD3 is predicted to fold into an N-terminal region unique to DALRD3 homologs and a C-terminal DALR anticodon binding domain (Figure 4G, N-terminal region in gray, DALR domain in violet). The C-terminal DALR domain of DALRD3 is predicted to form an all alpha-helical bundle that is characteristic of DALR domains found in arginyl tRNA synthetases and a subset of glycine tRNA synthetases^20,21^. Based upon the AlphaFold model, the R517 residue lies within the last alpha helix of the all alpha-helical bundle (Figure 4G, R517 residue in red). Moreover, the R517 residue lies on the surface of the DALR domain with the positively charged side chain partially exposed to solvent (Figure 4H). The R517C variant in the affected patient would change the amphipathic arginine residue to a nonpolar cysteine residue and reduce the overall size of the side chain. The change to a hydrophobic cysteine residue could alter the overall folding of the alpha-helical bundle that impacts interactions with METTL2 and tRNAs. Moreover, the change from a positively charged arginine side chain to the uncharged cysteine side chain could disrupt electrostatic or hydrogen bond interactions within DALRD3 itself or with METTL2 and arginine tRNAs. Thus, the R517C variant could alter the structure of the DALR domain leading to perturbations in its function in tRNA modification.

Altogether, we identify a novel pathogenic variant in DALRD3 that substantiates the link between DALRD3-dependent tRNA modification and proper neurological function. Moreover, these results uncover the molecular basis for the deficit in tRNA modifications caused by the DALRD3 missense variant. The patient with the DALRD3-R517C variant exhibits many of the phenotypes displayed by two previously characterized patients with a homozygous nonsense variant in DALRD3^16^. However, the onset of seizures in the patient described here was later in life compared to the two previously characterized patients. Moreover, the patient described here exhibited less severe immobility and speech deficiencies compared to the two patients with a homozygous nonsense variant. We also note that fibroblast cells from the patient with the R517C missense variant retain ∼25% of the m3C modification in arginine tRNAs while patient cells with the DALRD3 frameshift variant exhibit nearly complete loss of m3C modification in arginine tRNAs^16^. Thus, the reduced severity of clinical symptoms associated with the DALRD3-R517C variant is consistent with the partial loss-of-function molecular phenotype compared to the nearly complete loss of function in the DALRD3 frameshift variant. Our findings set the stage for determining the biological pathways dependent upon DALRD3-dependent tRNA modifications that could be modulated in patients with epileptic encephalopathy.

## Materials and Methods

Materials and Methods can be found in the Supplemental information file.

## Data Availability

All data produced in the present work are contained in the manuscript.

## Acknowledgements

We thank the patient and their family for participation in this study. We thank the Fu Lab for comments on this manuscript. This work was funded by NIH R01 GR532955 to D.F.

