## Supplemental Information for "Epileptic encephalopathy linked to a DALRD3 missense variant impaired in tRNA modification function"

### Whole-exome sequencing

The raw sequencing data was processed using our in-house developed pipeline, as described previously[1]. The resulting Vcf files were uploaded into the Cartagenia/Alissa clinical informatics platform (version 5.1.4, Agilent Technologies). Variant selection, analysis and classification were performed as previously described[2]. First, quality filtering of the called variants was performed, excluding all those with a read depth 0.1% in the population databases were excluded from further analysis as they are considered benign. After filtering, variants were evaluated for their potential pathogenicity using in silico prediction software tools that are part of the Alamut Batch software (version 2.12; Interactive Biosoftware, Rouen, France), including SIFT, PolyPhen-2, MutationTaster, AlignGVGD, PhyloP and Grantham distance and four different splice site prediction programs (NNsplice, MaxEntScan, GeneSplicer and SpliceSiteFinder-Like). Variants were classified as “likely benign” (LB, class 2), “variant of uncertain significance” (VUS, class 3), “likely pathogenic” (LP, class 4) or “pathogenic” (P, class 5), largely based on ACMG guidelines[3]. In addition, we searched for scientific literature (PubMed and databases) that report identical or similar gene variants, including the Human Genome Mutation Database (HGMD) and ClinVar, for known (likely) pathogenic variants reported in patients.

### Tissue culture

The DALRD3 R517C fibroblast cells were obtained by skin biopsy from the identified patient. The wildtype human fibroblast cell line was previously described and obtained from a healthy patient[4]. The 293T human embryonic cell line was obtained from ATCC (CRL-3216). Fibroblast and 293T cell lines were cultured in Dulbecco’s Minimal Essential Medium (DMEM) supplemented with 10% fetal bovine serum (FBS), 1X penicillin and streptomycin

(ThermoFisher), and 1X Glutamax (Gibco) at 37 °C with 5% CO2. Cells were passaged every 3 days with 0.25% Trypsin.

**Western blotting of patient cell samples**

Fibroblast cell lysates were prepared by resuspending cells in hypotonic lysis buffer (20 mM HEPES pH 7.9, 2 mM $MgCl_2$, 0.2 mM EGTA, 10% glycerol, 0.1 mM PMSF, 1 mM DTT) and incubation on ice for 5 minutes. The resuspended cells were subjected to three consecutive freeze-thaw cycles in liquid nitrogen and a 37 °C water bath. NaCl was added to the lysates to a final concentration of 0.4 M, incubated on ice for 5 minutes, and the lysates spun at 14,000 × g for 15 min at 4 °C. After centrifugation, the supernatant was collected and flash frozen in liquid nitrogen before storage at -80 °C.

Cellular extracts and purified protein samples were fractionated on NuPAGE Bis-Tris polyacrylamide gels (Thermo Scientific) followed by transfer to Immobilon FL PVDF membrane (Millipore) for immunoblotting. Antibodies were against the following proteins : DALRD3 (MA1-21315, Thermo Fisher), FLAG (NC9261069, Thermo Fisher), and actin (CST). Primary antibodies were detected using IRDye 800CW Goat anti-Mouse IgG (SA5-35521, Thermo Fisher) or Rabbit (SA5-35571, Thermo Fisher) or IRDye 680RD Goat anti-Mouse IgG (926-68070, LI-COR Biosciences) or goat anti-Rabbit IgG (925-68071). Immunoblots were scanned using direct infrared fluorescence via the Odyssey System (LI-COR Biosciences).

**Primer extension analysis**

Total RNA from human cells was extracted and purified using TRIzol LS reagent (Thermo Fisher). Primer extension was performed as previously described[5]. Briefly, 3 μg of total RNA was pre-annealed with a 5′-$^{32}$P-radiolabelled oligonucleotide complementary to tRNA-Arg-UCU or CCU. The mixture was heated at 95 °C for 3 minutes followed by slow cooling to 45 °C. After cooling, 14 μl of extension mix (0.12 μl of avian myeloblastosis virus reverse transcriptase

(Promega), 2.8 µl 5X RT buffer, 1.12 µl 1 mM dNTPs, RNase-free water to 14 µl) was added to each reaction and incubated at 45 °C for 1 hour. Samples were mixed with 2x formamide denaturing dye, heated to 95 °C for 3 minutes and resolved on an 20% polyacrylamide, 7 M urea, 1xTBE denaturing gel. Phosphorimaging was performed on an Azure Sapphire phosphorimager and analyzed using NIH ImageJ software.

**Transient transfections and protein-RNA purifications**.

293T cells were transfected via calcium phosphate transfection method. Briefly, $2 \times 10^6$ cells were seeded on 100 × 20 mm tissue culture grade plates (Corning) followed by transfection with 20 µg of plasmid DNA. After 48 hours post-transfection, cells were dissociated from the plate using trypsin, harvested at 700 × g for 5 minutes followed by washing with PBS. Cell pellets were resuspended in 0.5 mL of hypotonic lysis buffer (20 mM HEPES pH 7.9, 2 mM MgCl2, 0.2 mM EGTA, 10% glycerol, 0.1 mM PMSF, 1 mM DTT). Cells were kept on ice for 5 minutes followed by three freeze-thaw cycles in liquid nitrogen and a 37 °C water bath. NaCl was added to the lysates to a final concentration of 0.4 M, incubated on ice for 5 minutes and the lysates spun at 14,000 × g for 15 min at 4 °C. After centrifugation, 500 µl of the lysate was removed and diluted with an equal volume of hypotonic lysis buffer containing 0.2% NP-40. Cell lysates were flash frozen in liquid nitrogen and stored at -80 °C.

FLAG-tagged proteins were purified by incubating whole cell lysates with 50 µl of MonoRab Anti-DYKDDDDK Magnetic Beads (GenScript) for two hours at 4 °C. Magnetic resin was washed three times with 1X HLB200 (20 mM HEPES pH 7.9, 2 mM $MgCl_2$, 0.2 mM EGTA, 10% glycerol, 0.1% NP-40, 200 mM NaCl, 0.1 mM PMSF, 1 mM DTT). Proteins were eluted by resuspending resin in 1X NuPAGE LDS Sample Buffer supplemented with 50 mM DTT and heating at 95 °C for 5 minutes. Input lysates and purified proteins were fractionated on a NuPAGE Bis-Tris PAGE (ThermoFisher) and transferred to Immobilon-FL PVDF Membrane (Millipore Sigma). Immunoblotting was performed as described above.

For RNA copurification analysis, the resin was resuspended in 100 µL of Molecular Biology Grade RNAse-free water (Corning). 15 µl of the resuspended resin was taken for immunoblotting analysis where the beads were mixed with 5 µl of 4X NuPAGE LDS Sample Buffer supplemented with 50 mM DTT and heating at 95 °C for 5 minutes prior to loading onto a NuPAGE Bis-Tris PAGE (ThermoFisher). RNA extraction followed the TRIzol LS RNA extraction protocol (Invitrogen). Recovery control RNA (20 ng) was added to the purified bead samples. RNA was resuspended in 10 µl of RNAse-free water and loaded onto a 10% polyacrylamide, 7 M urea gel. The gel was then stained with SYBR Gold nucleic acid stain (Invitrogen) to visualize RNA.

**Ethics approval and consent to participate**

IRB of University Medical Centre of Groningen gave ethical approval for this work. The patient and family provided consent for participation in the study and publication of the results.

**Availability of data and materials**

All data and materials are available upon request.

A

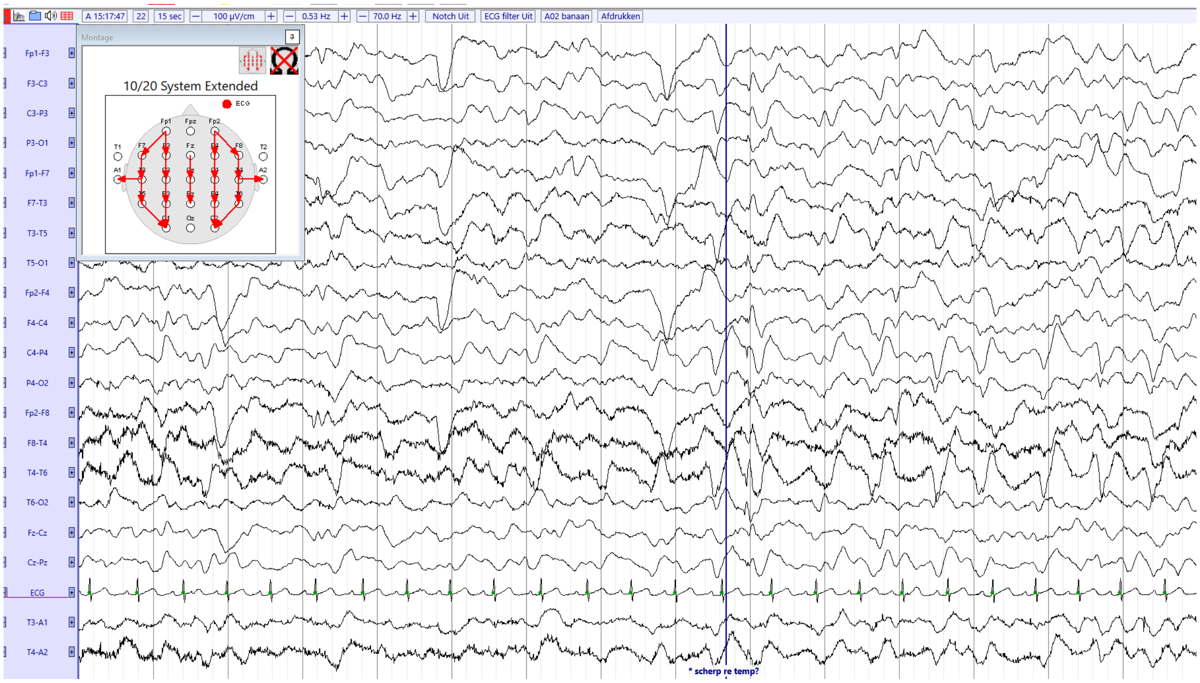


B

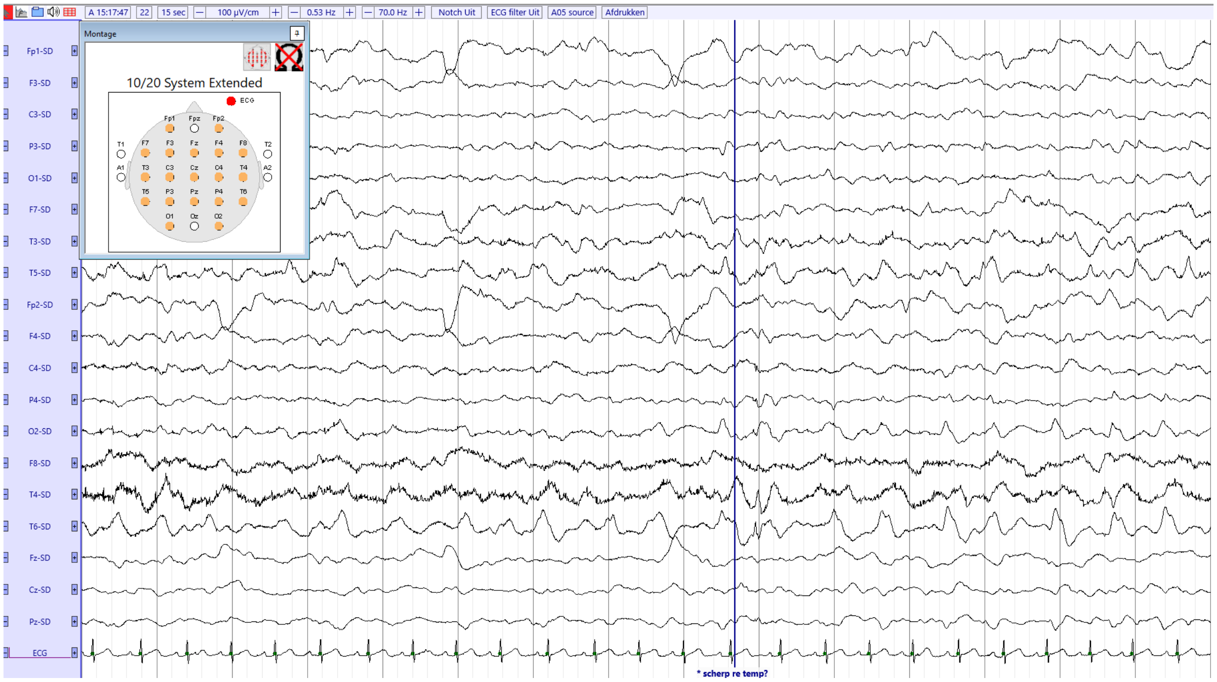


**Supplemental Figure 1.** Epileptiform discharges detected at the right temporal electrode during EEG testing of the patient in this study. (A) Bipolar and (B) Laplacian montages.

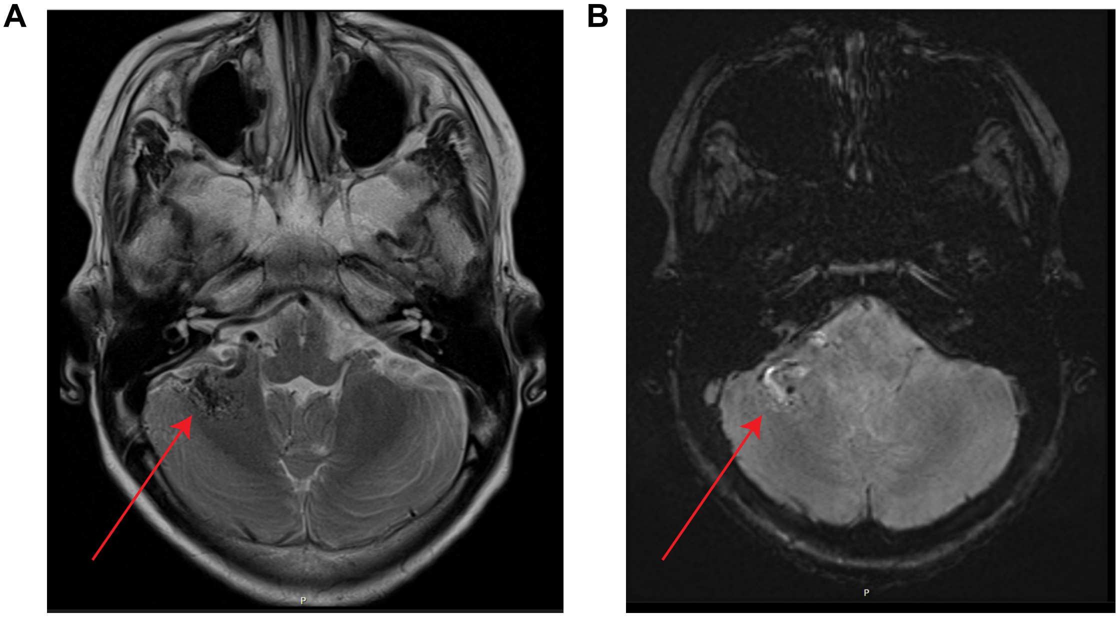


**Supplemental Figure 2.** Right cerebellar arteriovenous malformation detected in the patient in this study. (A) Transversal T2 TSE weighted MRI scan. (B) Transversal SWI (susceptibility weighted) scan.

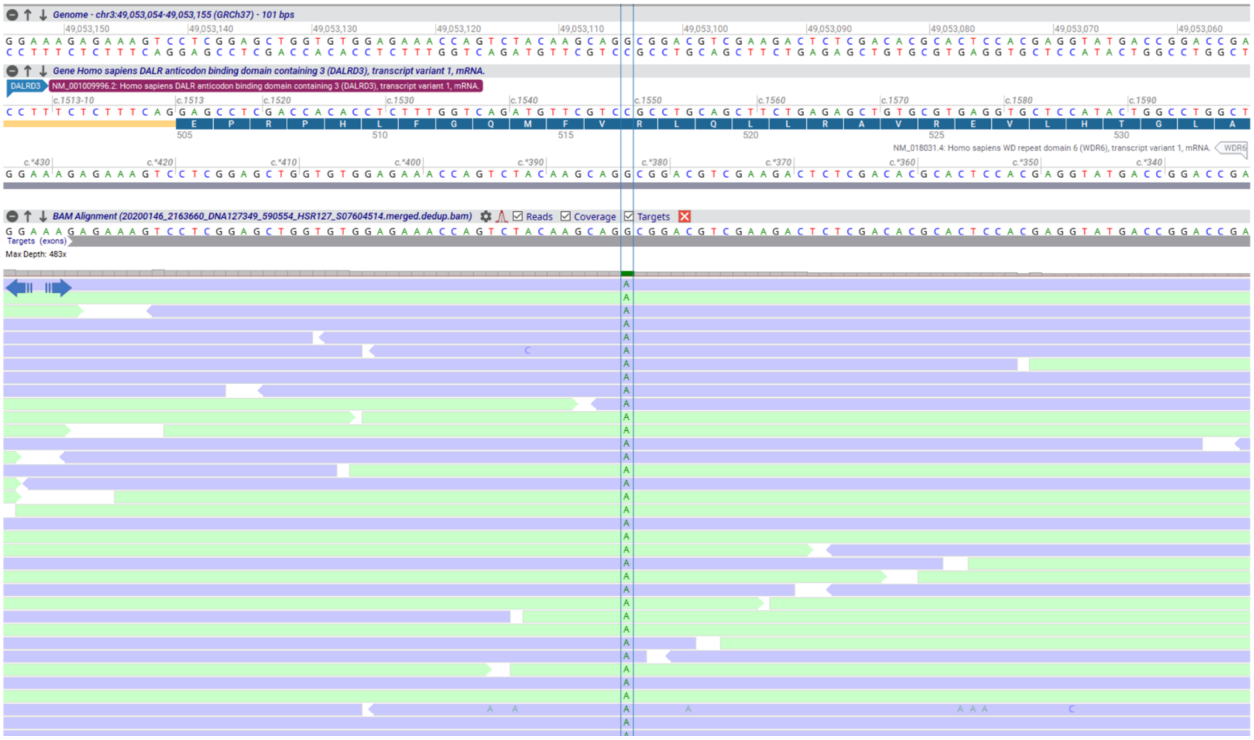

**Supplemental Figure 3.** Identification of a homozygous missense variant in the *DALRD3* gene of the patient in this study. Whole exome sequence analysis output.

**A**

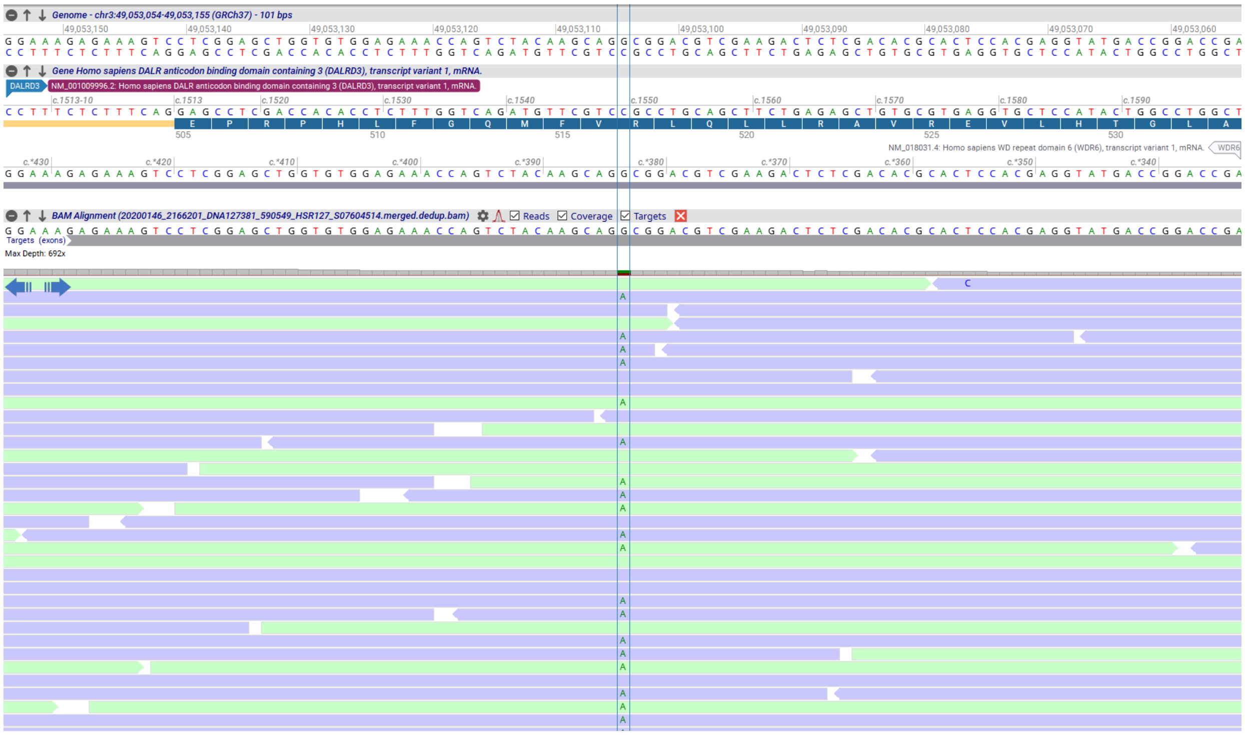

**B**

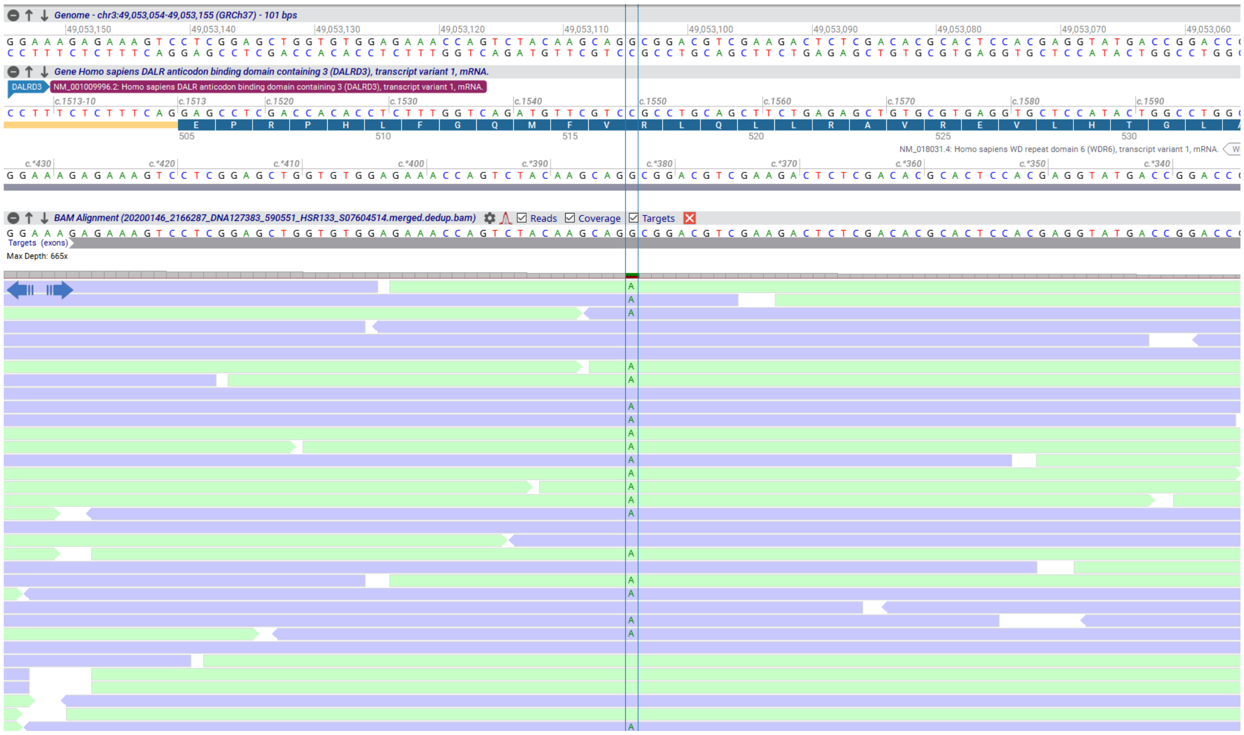

**Supplemental Figure 4.** The parents of the affected patient in this study are heterozygous carriers of the *DALRD3* missense variant. Whole exome sequence analysis of (A) Parent 1 and (B) Parent 2.
